# Obstructive sleep apnea is highly prevalent in COVID19 moderate to severe ARDS survivors: Findings of level I Polysomnography in a tertiary care hospital

**DOI:** 10.1101/2021.05.04.21256593

**Authors:** Abhishek Goyal, Khushboo Saxena, Avishek Kar, Alkesh Khurana, Parneet Kaur Bhagtana, Chinta Siva Koti Rupa Sridevi, Abhijit Pakhare

## Abstract

**Study Objectives:** Studies have found Obstructive Sleep Apnea (OSA) as a risk factor for increased risk for COVID19 Acute respiratory Distress Syndrome (ARDS); but most of the studies were done in already known patients of OSA. This study was done to find prevalence of OSA in patients with COVID-19 related acute respiratory distress syndrome.

**Methodology:** A hospital based longitudinal study was conducted among COVID 19 Intensive Care Unit (ICU) survivors. All consecutive COVID19 with moderate to severe ARDS were evaluated for OSA by Level I Polysomnography (PSG) after 4-6 weeks of discharge. Prevalence of OSA and PSG variables {Total sleep time, Sleep efficiency, sleep stage percentage, Apnea Hypopnea Index (AHI), T90, nadir oxygen} was estimated.

**Results:** Out of 103 patients discharged from ICU during study period (October 2020 to 15 December 2020), 67 underwent Level I PSG. Mean Age was 52.6±10.9 years and mean Body Mass Index was 27.5 ± 6.2 Kg/m^2^. Total sleep time was 343.2 ± 86 minutes, sleep efficiency was 75.9±14.2%. OSA (AHI ≥5) was seen in 65/67 patients and 49 patients had moderate to severe OSA (i.e. AHI ≥ 15).

**Conclusion:** Moderate-severe OSA was highly prevalent (73%) in COVID19 moderate to severe ARDS survivors. Role of OSA in pathophysiology of COVID19 ARDS needs further evaluation.

**Highlights:**

- This study was done to find prevalence of OSA in patients with COVID-19 related Acute respiratory distress syndrome
- Moderate-severe OSA is highly prevalent (73%) in COVID19 ARDS survivors.
- To the best of our knowledge, it is first study in which level I PSG was done in COVID19 survivors.

## Introduction

COVID-19 has been one of the biggest catastrophe in the history of mankind Till date 100 million people have been infected and more than 2 million have died of this infection.

Severity of COVID-19 infection has been attributed to several comorbid conditions like systemic hypertension, diabetes mellitus, coronary artery disease and chronic kidney disease.^1^ In an attempt to decipher the pathophysiology and predisposing conditions, researchers are exploring newer avenues. Published articles have also assessed Obstructive Sleep Apnea(OSA) as a probable risk factor for severe COVID-19 disease due to shared pathophysiological pathways.^2,3^A retrospective study which was based on electronic health record has identified OSA as risk factor for mortality in COVID19 ARDS.^4^ Moreover the relationship of severity of OSA with that of COVID-19 has not been precisely documented.

This study was done to find prevalence of OSA in patients with COVID-19 related moderate to severe Acute respiratory distress syndrome.

## METHODOLOGY

This is a single centred prospective study conducted at a tertiary care teaching hospital in India. This study was carried over a period of two months (October 2020 to December 2020) on consecutive symptomatic COVID19 related acute respiratory distress syndrome patients who were discharged from ICU or isolation wards.

Aim of this study was to find prevalence of OSA in patients with COVID-19 related Acute respiratory distress syndrome.

### Inclusion Criteria

Patient meeting all the criteria were included

1. Age ≥ 18 years.
2. Patients positive for COVID 19 by RT PCR of nasopharyngeal and oropharyngeal swabs at the time of ICU admission
3. Patients with P/F <200 and fulfils criteria of ARDS.
4. Patients must not require oxygen support at time of sleep study.
5. Patients must have tested negative COVID 19 (nasopharyngeal swab-RT PCR) at least 3 weeks prior to study

### Exclusion criteria

1. Patients who refused to give consent for PSG and participation in study
2. Patients who were not able to sit and stand without support even after 3 weeks of discharge.
3. Patients who required oxygen support after 3 weeks of discharge from ICU.
4. Any symptoms of active pulmonary infection.

All consecutive patients who were getting discharged from ICU were screened for general condition, maximum oxygen requirement during hospital course and interface of oxygen supplementation were noted. Appointment for polysomnography was allotted after 3 weeks of negative nasopharyngeal and oropharyngeal COVID19 RT PCR report.

On the day of PSG, they were again screened for any symptoms suggestive of respiratory infection, fever, throat pain, dyspnea and oxygen requirement prior to sleep study appointment. Patients were enrolled when found fit for study as per inclusion criteria. All demographic parameters, co morbidities, anthropometric parameters, STOP BANG, NoSAS, Berlin questionnaire (BQ) and Epworth Sleepiness Scores (ESS) were recorded prior to study. Questions for OSA and sleep disorders were filled for the period before subjects acquired COVID19.

From ICU records, weight of patient on ICU admission was noted. Weight was again taken on the day of PSG. Weight loss or gain from time of ICU admission was noted for each patient.

Precautions taken during Polysomnography: Following precautions were taken during PSG in all patients to prevent spread of infection among patients and staff:

1. No relatives were allowed to sleep in sleep lab room. Only patient was admitted in sleep lab.
2. Every patient was again checked for temperature, COVID-19 symptoms by sleep technician on day of sleep study.
3. Team of sleep technicians were assigned as per rotation protocol. Technicians had to wear personal protective equipment as per hospital protocol which included disposable eye cover, N-95 respirator, face shield, gown, gloves and shoe covers.
4. Disposable bed covers were used which were discarded after each study.
5. Each sleep room was properly sanitized as per hospital protocol. After PSG, room was fumigated and locked for 72 hours for safety purposes.
6. Level I PSG equipment were sanitized after every sleep study and were not used for next 72 hours.

## Polysomnography (PSG)

All patients underwent level I PSG (Philips Respironics Alice 6). The following parameters were monitored during PSG: Electroencephalogram (EEG): frontal, central and occipital, electro-oculogram (EOG), sub-mentalis Electromyogram (EMG), nasal and oral airflow, anterior tibialis EMG, body position and electrocardiogram. Additionally, thoracic and abdominal movements were recorded by inductance plethysmography (zRIP). Oxygen saturation (SpO2) was monitored using a pulse oximeter. The tracing was scored using 30 second epochs. Scoring of events was done according to AASM scoring manual version 2012.^5^Apneas were marked when there is drop in peak signal excursion by ≥ 90% of pre event baseline with duration of the ≥ 90% drop in sensor signal ≥ 10 seconds. Hypopneas were scored if peak signal excursions drop by ≥ 30% of pre event baseline with duration of the ≥ 30% drop in signal excursion was ≥ 10 seconds with associated ≥ 3% oxygen desaturation from pre event baseline or event was associated with an arousal. The severity of obstructive sleep apnea was described by number of apneas and hypopneas per hour of sleep (apnea-hypopnea index [AHI]). OSA was defined as an AHI ≥5. OSA into three categories on the basis of AHI: mild (5-14.9) to moderate (AHI 15-29.9) and severe (AHI ≥30).

### Ethics review

Approval for ethics approval was taken approval from AIIMS Bhopal Institutional Human Ethics committee (IHEC-LOP/2020/IM309). Patients were counselled regarding purpose of the study and consent was taken from every patient.

### Data analysis

Data analysis was done using R software^6^ and *gtsummary*^7^ packages. We have summarized nominal variables with count and percentage and numerical variables with mean and its standard deviation. We have stratified our data across those who required oxygen and those who required ventilatory support (NIV/HFNC/IMV). Then we have tested difference in distribution of numerical and categorical variables across these groups by using t-test and Chi-square test respectively. p-value less than 0.05 was considered as statistically significant.

## Results

Consecutive patients of COVID ARDS admitted in ICU were recruited after 3-4 weeks of discharge. Out of 103 patients discharged during study period (October 2020 to 15 December 2020), 67 patients (46 male, 21 female) underwent Level I PSG.(Figure 1)

**Figure 1:**
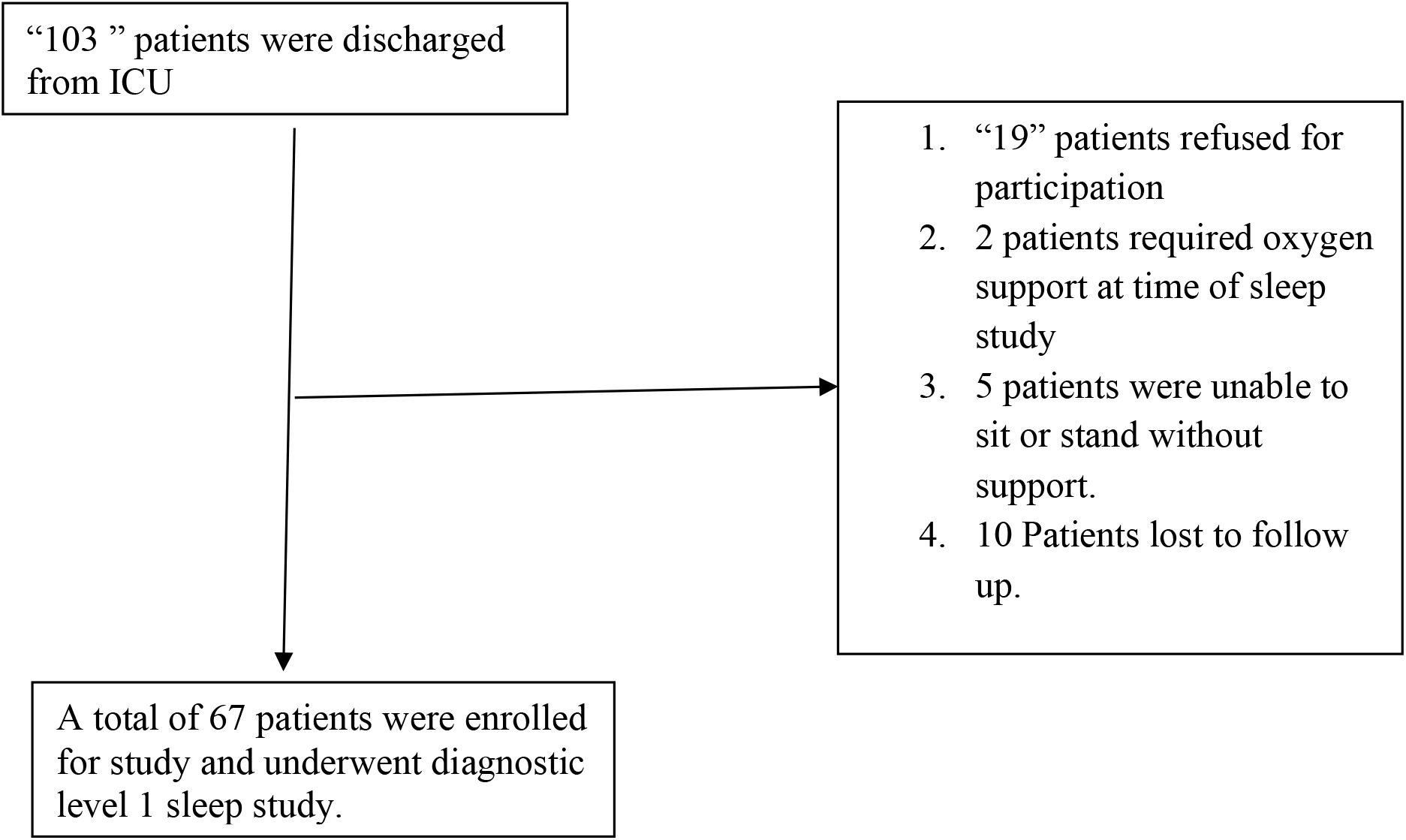
Flow chart of enrolment of patients for current study

Mean Age was 52.6± 10.9 years and mean BMI at time of PSG was 27.5± 6.2 Kg/m^2^. (Table 1)Seventeen out of sixty seven had BMI≥30 Kg/m^2^and 50 had BMI<30 Kg/m^2^.

**Table-1.**
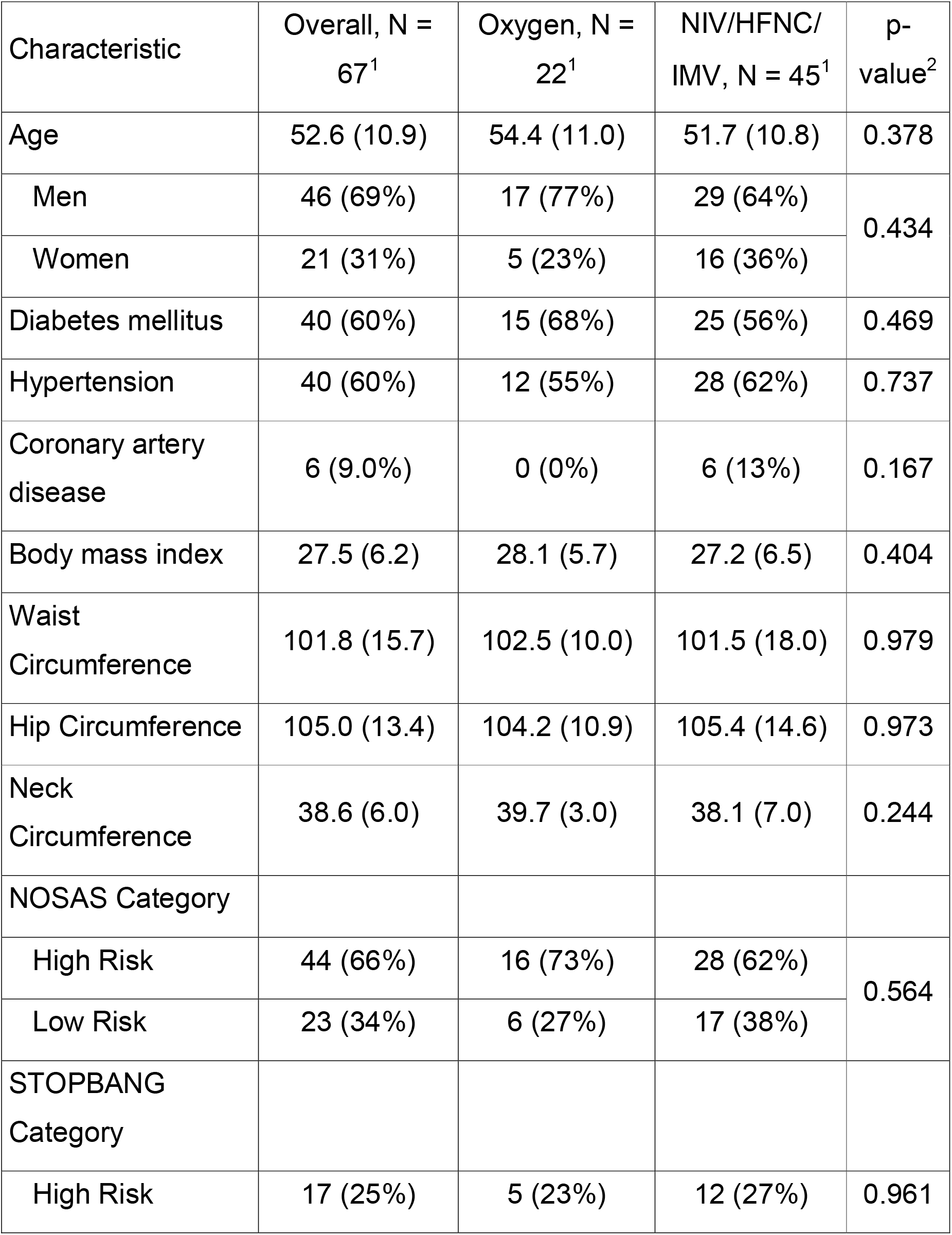
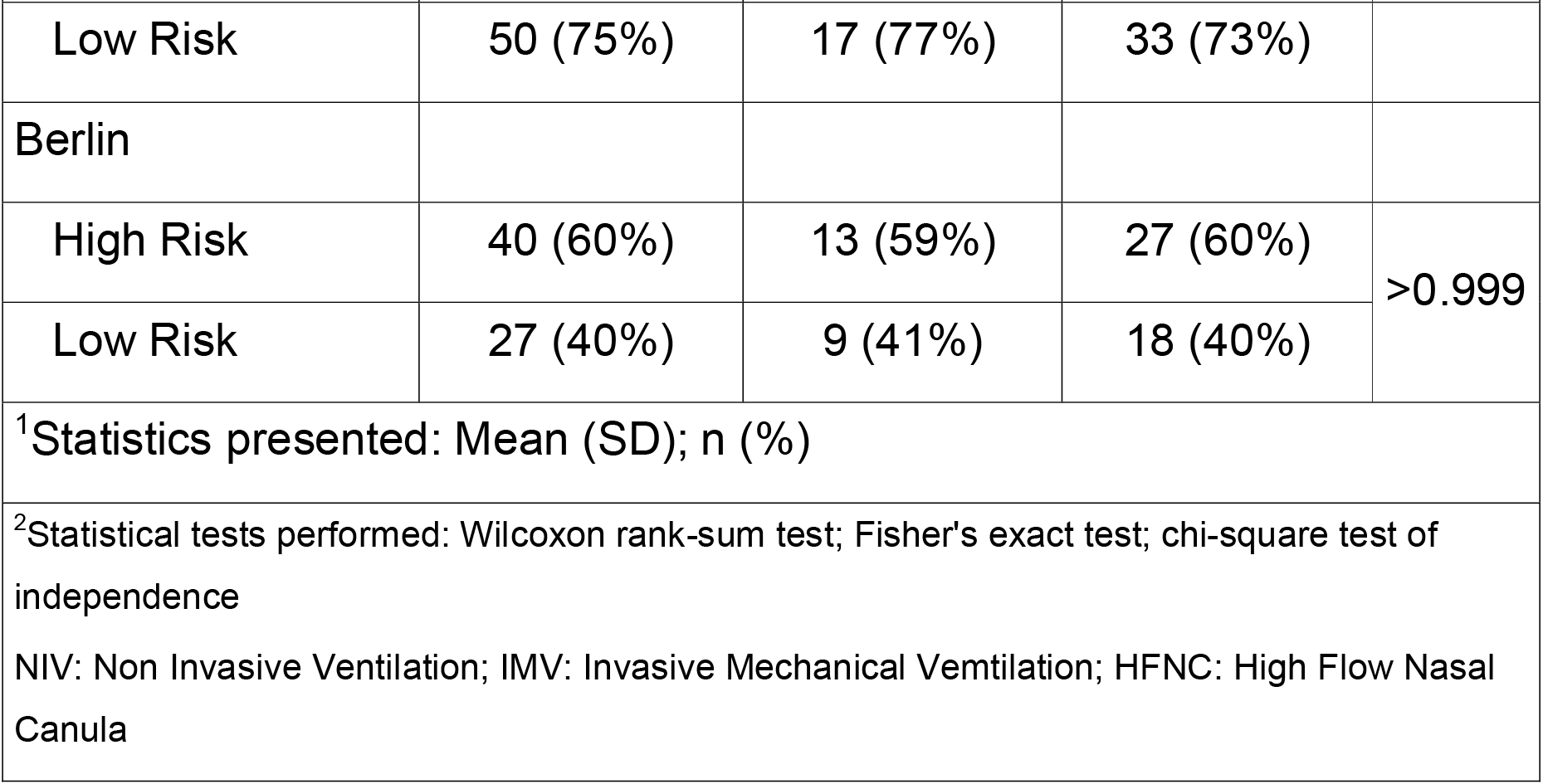
Baseline demographic and clinical characteristics.

Median weight loss of patients was 0.0 Kg (IQR 0.0-4.2) from time of ICU admission and PSG. Six patients had weight gain and 29 had weight loss; rest 32 patients had constant weight (i.e.less than 1 kg weight difference) between these two timelines.

Twenty two patients required oxygen varying from 5 LPM to 15 LPM during their ICU stay and did not require either Non Invasive Ventilation(NIV) or Invasive Mechanical Ventilation (IMV). Thirty two patients could be successfully managed with either NIV(20) or High Flow Nasal Canula (12) during ICU stay and 13 patients had required IMV. History of Diabetes Mellitus (DM) and hypertension was present in 40 patients each.

### PSG characteristics

(Table 2): Total sleep time was 343.2± 86 minutes. Sleep efficiency was 75.9±14.2%. Mean time (%) during N1, N2, N3 and Rapid Eye movement (REM) was 15.9%, 58.7%, 8.5% and 19% percentage respectively. Mean AHI was 29.1± 21.9 per hour and arousal index was 23.9±13.3 OSA (defined by AHI≥5) was seen in 65/67 patients and 49 patients had moderate to severe OSA (i.e. AHI was more than 15).

**Table-2.** Distribution of PSG variables.

| Characteristic | Overall, n = 67<sup>1</sup> | Oxygen, n=22<sup>1</sup> | NIV/IMV, n = 45<sup>1</sup> | p-value<sup>2</sup> |
| --- | --- | --- | --- | --- |
| Total sleep time (in minutes) | 343.2 (86.0) | 358.5 (53.3) | 335.7 (97.8) | 0.580 |
| Sleep efficiency (%) | 75.9 (14.2) | 78.3 (8.4) | 74.7 (16.2) | 0.873 |
| Sleep latency (in minutes) | 19.8 (22.0) | 17.3 (21.4) | 21.1 (22.5) | 0.378 |
| N1 in percentage | 15.9 (10.1) | 11.9 (4.5) | 17.9 (11.4) | 0.031 |
| N2 in percentage | 58.7 (12.4) | 60.4 (10.7) | 57.9 (13.2) | 0.522 |
| N3 in percentage | 8.5 (7.6) | 10.5 (9.3) | 7.6 (6.5) | 0.268 |
| REM in percentage | 19.0 (14.8) | 19.0 (8.4) | 19.0 (17.2) | 0.419 |
| AHI (Apnea Hypopnea Index) | 29.1 (21.9) | 24.9 (20.2) | 31.1 (22.6) | 0.175 |
| NREM AHI | 28.2 (22.9) | 23.1 (19.8) | 30.7 (24.1) | 0.184 |
| REM AHI | 31.2 (25.0) | 26.3 (22.4) | 33.7 (26.1) | 0.207 |
| OSA severity |  |  |  |  |
| No OSA | 2 (3.0%) | 1 (4.5%) | 1 (2.2%) | 0.395 |
| Mild | 16 (24%) | 6 (27%) | 10 (22%) |  |
| Moderate | 25 (37%) | 10 (45%) | 15 (33%) |  |
| Severe | 24 (36%) | 5 (23%) | 19 (42%) |  |
| Positional OSA | 33 (49%) | 12 (55%) | 21 (47%) | 0.730 |
| NREM Predominant | 13 (19%) | 3 (14%) | 10 (22%) | 0.765 |
| REM Predominant | 14 (21%) | 5 (23%) | 9 (20%) |  |
| Arousal index | 23.9 (13.3) | 23.2 (13.7) | 24.3 (13.2) | 0.694 |
| ODI | 22.8 (26.9) | 15.3 (17.6) | 26.5 (30.0) | 0.055 |
| T90 | 6.0 (17.4) | 0.6 (1.3) | 8.7 (20.7) | 0.049 |
| Weight loss (in Kg) | 3.1 (5.5) | 2.4 (4.7) | 3.5 (5.8) | 0.654 |
| <sup>1</sup>Statistics presented: Mean (SD); n (%) |  |  |  |  |
| <sup>2</sup>Statistical tests performed: Wilcoxon rank-sum test; Fisher's exact test; chi-square test of independence |  |  |  |  |
| AHI: Apnea Hypopnea Index; NREM: Non-Rapid Eye Movement; ODI: Oxygen Desaturation Index; OSA: Obstructive Sleep Apnea; REM: Rapid Eye Movement; T90: Percentage sleep time spent with spO2<90% |  |  |  |  |

## Discussion

This study shows that OSA is highly prevalent in patients with COVID19 induced ARDS. To the best of our knowledge, it is first study in which level I PSG was done in COVID19 survivors.

Breathing disruption seen in OSA is associated with sympathetic activity surge and hypoxemia which predisposes patients to cardiovascular and neurological complications.^8,9,10^ OSA has been associated with obesity, hypertension, Diabetes Mellitus (DM), Coronary artery disease, arrythmia, nocturia, chronic kidney disease, dyslipidaemia, metabolic syndrome, and pulmonary embolism.^8,11,12^ Association of COVID 19 ARDS is now known with various risk factors like obesity, older age, male sex, DM, hypertension, Coronary Artery Disease and Chronic Kidney Disease.^13^ Most important of these factors were age, obesity, and DM; all these three factors are closely interlinked with OSA. SARS CoV2 enters cells through Angiotensin Converting Enzyme-2 receptors in lungs and there is increased expression of these receptors in obese individuals.^14^

OSA might precipitate inflammation caused due to COVID19 by multiple pathways: OSA itself leads to hypoxemia and hypoventilation during sleep and superimposed COVID19 can lead to profound hypoxemia.^15^ Studies have shown increased inflammatory markers (like IL6 and CRP) in patients with OSA.^16,17^ OSA is a known risk factor for cardiac complications (hypertension, heart failure, acute cardiovascular events & arrhythmias) and thus may increase cardiac morbidity and mortality in patients when they contract COVID infection.^8,11,12^ DVT and PE risks were shown to be 3.50- and 3.97-fold higher respectively in OSA cohort than in the reference cohort.^18^ Hemodynamic alterations in OSA lead to polycythaemia and sluggish blood flow, which can possibly lead to procoagulant state.^19,20,21^ This hypercoagulable state can probably further increase risk for COVID related coagulopathy.

Recently few studies have tried to find out possibility of poor outcome in COVID19 with pre-existing OSA. In an electronic health record review, mortality was found to be much higher in patients with known OSA (11.7%) compared to controls (6.9%) with an odds ratio of 1.7923.^4^ In the CORONADO study, 144/1189 patients were already known case of OSA and presence of OSA was associated with poor outcome in COVID 19 related illness.^22^In our previous study involving 213 patients, screening questionnaires for OSA {STOPBANG, Berlin Questionnaire (BQ), NoSAS} were more likely to be positive in patients who died compared to patients who survived.^23^ Proportion of patients who were classified as high risk for OSA by various OSA screening tools significantly increased with increasing respiratory support (p<0.001 for STOPBANG, BQ, ESS and p=0.004 for NoSAS). On binary logistic regression analysis, only two factors: age≥ 55 and STOPBANG score ≥5 (correlating with moderate-severe OSA) were found to independent effect on mortality.

In all previous done work, association with severity of COVID19 was tried to be found in already known patients of OSA. But it is a known fact that more than 90% of patients with OSA remain undiagnosed.^24^ None of patients (in current study) were ever evaluated for OSA before COVID19 infection and yet 73% of them had moderate-severe OSA.

In pre-COVID19 era, very few studies were done to evaluate sleep disorders (with level I PSG) in ARDS survivors. In a study done from India, PSG was done in 20 ARDS survivors after 4 weeks of discharge from ICU: 2 had Central Sleep Apnea and one had OSA.^25^In another study from United Kingdom, level I PSG was done immediately after discharge from ICU; out of 15 mechanically ventilated patients, 11 had OSA (AHI≥5) and 9 (60%) had Moderate-severe OSA (AHI≥15).^26^In another study, seven ARDS survivors without previous sleep complaints reported difficulty sleeping 6 months after discharge from hospital.^27^ All these seven patients underwent PSG and only one had apnea. Further, N3 and REM sleep was significantly reduced, which was consistent with previous reports.

The high prevalence of OSA seen in patients with COVID19 ARDS in our study hints that OSA might be an independent risk factor for poor prognosis in patients with COVID19. To prove this however, PSG should be done in asymptomatic or minimally symptomatic individuals diagnosed with COVID19.

Untreated OSA can lead to cardiac complications and persistent symptoms including fatigability, insomnia, which are commonly seen in followup of COVID`19 patients.^28^ So in our opinion OSA should be ruled out in COVID19 ARDS survivors with these complaints and if moderate-severe OSA is present, it should be treated with Continuous Positive Airway Pressure (CPAP) therapy.^29,30^

Most important strength of this study is that gold standard level I PSG was done in consecutive ICU survivors. To the best of our knowledge, this is the first study in which PSG was done in COVID19 patients.

Important limitations of this study were:1) Presence of OSA could not be determined in patients who died during hospitalisation. This may have resulted in underestimation of prevalence of moderate to severe OSA. Although our previous work have shown increased mortality among those with high risk for OSA (as assessed with OSA screening questionnaires).^23^

2) Association between presence of OSA and severity of COVID19 could not be explored owing to inclusion of participants from only ICU. For this, patients with minimal or mild symptoms of COVID19 should have been included in study. However due to resource constraints, we restricted our study to ICU survivors. Participants of this study were patients from ICU only therefore findings should not be generalised to all COVID19 patients.

To summarise, moderate-severe OSA is highly prevalent (73%) in COVID19 moderate to severe ARDS survivors and it will be interesting to explore further effect of role of OSA in pathophysiology of COVID19 ARDS.

## Data Availability

Data will be made available if required by any reviewer.

## Acknowledgements

We want to thank Mr Amit Sen, Mr Chetan, Mr Ranjit and Ms Artee for performing polysomnography.

## Author Contribution

AG: Conceptualized, data collection, analysis, patient management and manuscript writing.

KS: Data collection, Patient management, manuscript writing

Av K: Patient management, Data collection, manuscript writing

AK: Patient management, Manuscript writing

PKB: Data collection, manuscript writing

CSKRS: Data collection, manuscript writing

AP: Analysis, manuscript writing

## Compliance with Ethical Standards

This is a retrospective analysis study and waiver of this study was taken from ethical clearance committee.

## Ethical approval

All procedures performed in studies involving human participants were in accordance with the ethical standards of the institutional and/or national research committee and with the 1964 Helsinki declaration and its later amendments or comparable ethical standards.

## Informed consent

Informed consent was obtained from all individual participants included in the study.

## OSA COVID Analysis (PSG)

